# Controlled Human Malaria Infection reveals that the Dantu blood group variant provides high level protection against uncomplicated malaria

**DOI:** 10.1101/2022.09.21.22280031

**Authors:** Silvia N Kariuki, Alexander W Macharia, Johnstone Makale, Wilfred Nyamu, Stephen L Hoffman, Melissa C Kapulu, Philip Bejon, Julian C Rayner, Thomas N Williams, the CHMI-SIKA Study Team

## Abstract

**Introduction:** The long co-evolution of *Homo sapiens* and *Plasmodium falciparum* has resulted in the selection of numerous human genetic variants that confer an advantage against severe malaria and death. One such variant is the Dantu blood group antigen, which is associated with 74% protection against severe and complicated *P. falciparum* malaria infections in homozygous individuals, similar to that provided by the sickle haemoglobin allele (HbS). Recent *in vitro* studies suggest that Dantu exerts this protection by increasing the surface tension of red blood cells, thereby impeding the ability of *P. falciparum* merozoites to invade them and reducing parasite multiplication. However, no studies have yet explored this hypothesis *in vivo*.

**Methods:** We investigated the effect of Dantu on early phase *P. falciparum* (Pf) infections in a controlled human malaria infection (CHMI) study. 141 sickle negative Kenyan adults were inoculated with 3.2×10^3^ aseptic, purified, cryopreserved Pf sporozoites (PfSPZ Challenge) then monitored for blood-stage parasitaemia for 21 days by quantitative polymerase chain reaction (qPCR) analysis of the 18S ribosomal RNA *P. falciparum* gene. The primary endpoint was blood-stage *P. falciparum* parasitaemia of ≥500/μl while the secondary endpoint was the receipt of antimalarial treatment in the presence of parasitaemia of any density. On study completion, all participants were genotyped both for Dantu and for four other polymorphisms that are associated with protection against severe falciparum malaria: α^+^-thalassaemia, blood group O, G6PD deficiency, and the rs4951074 allele in the red cell calcium transporter *ATP2B4*.

**Results:** The primary endpoint was reached in 25/111 (22.5%) non-Dantu subjects, in comparison to 0/27 (0%) Dantu heterozygotes and 0/3 (0.0%) Dantu homozygotes (p=0.01). Similarly, 49/111 (44.1%) non-Dantu subjects reached the secondary endpoint in comparison to only 7/27 (25.9%) and none of the 3 (0.0%) Dantu heterozygotes and homozygotes respectively (P=0.021). No significant impacts on either outcome were seen for any of the other variants under study.

**Conclusion:** This study reveals, for the first time, that the Dantu blood group is associated with high level protection against early, non-clinical, *P. falciparum* malaria infections *in vivo*. Learning more about the mechanisms involved could potentially lead to new approaches to the prevention or treatment of the disease. Our study illustrates the power of CHMI with PfSPZ Challenge for directly testing the protective impact of genotypes previously identified using other methods.

## Introduction

*Plasmodium falciparum* malaria has been the pre-eminent cause of child morbidity and mortality in the tropics and sub-tropics for much of the last 5000 years. As a consequence, it has had a substantial impact on the human genome through the positive selection of multiple polymorphisms that confer a survival advantage against the disease [1]. The best studied affect the biology of red blood cells (RBCs), which host malaria parasites for most of their life cycle in humans, the rs334 A>T β^s^ sickle mutation in *HBB* [2], α-thalassaemia [3] and blood group O [4] all being important examples.

Recently, we identified a new variant which is associated with high-level protection against severe *P. falciparum* malaria to a degree that is close to that of sickle cell trait (HbAS), the strongest malaria-protective condition yet described [5]. The rare Dantu blood group antigen, which results from a genetic rearrangement within the glycophorin (*GYP*) cluster, was shown to confer 74% protection against severe malaria in homozygous individuals [6, 7]. Subsequent *in vitro* studies have suggested that this protection is explained by the resistance of Dantu RBCs to invasion by *P. falciparum* merozoites [8], thereby preventing infections from progressing to become severe or ultimately fatal. However, this hypothesis has not been tested directly *in vivo* to date.

In the current study, we have investigated the impact of the Dantu blood group on *in vivo P. falciparum* parasite growth and clinical disease progression through a controlled human malaria infection (CHMI) study with aseptic, purified, cryopreserved *P. falciparum* sporozoites (PfSPZ Challenge) conducted in semi-immune Kenyan adults. To the best of our knowledge, this is the first time that CHMI has been used to directly explore the impact of Dantu on parasite growth *in vivo*.

## Methods

### Study design and population

The primary aim of the Kenya CHMI study was to investigate the impacts of naturally acquired immunity on early-phase malaria infections [9, 10]. Briefly, 161 healthy adult volunteers living in areas of varying malaria transmission were inoculated by direct venous inoculation (DVI) with 3.2×10^3^ *P. falciparum* sporozoites (PfSPZ) of Sanaria® PfSPZ Challenge (NF54) [11, 12]. Because of its major impacts on both malaria susceptibility [13] and disease progression [14], and in view of results from a previous CHMI with PfSPZ Challenge conducted in Gabon [15], recruitment was restricted to those who were negative for both sickle cell trait and disease. Venous blood samples were collected twice daily from days 7 to 14, and then once every day from day 15 until the end of the experiment on day 21, and screened for parasitaemia by quantitative polymerase chain reaction (qPCR) analysis of the *P. falciparum* 18S ribosomal RNA gene. For each participant the endpoint was considered met, and anti-malarial treatment administered, when a threshold of 500 *P. falciparum* parasites/µl was reached. Participants were treated earlier if signs and symptoms were observed in association with blood film positivity at any parasite density, and on day 21 post-inoculation regardless of outcome. Participants were recruited during 2016, 2017 and 2018 into three successive cohorts from three different malaria transmission zones: Kilifi North (no-to low-transmission) and Kilifi South (moderate transmission), both on the coast, and Ahero (moderate to high transmission) in Western Kenya. The study was conducted at the KEMRI-Wellcome Trust Research Programme in Kilifi, Kenya and was registered on ClinicalTrials.gov (NCT02739763).

### Genotyping for Dantu and other malaria protective variants

Whole blood samples were collected at the point of recruitment into tubes containing EDTA and stored at -80^0^C pending batch processing at the end of the study. Genomic DNA was extracted following the manufacturer’s instructions using a QIAmp 96 DNA QIAcube HT kit on a QIAcube HT System (QIAGEN, Manchester UK). Genotyping for Dantu was performed using ABI TaqMan SNP genotyping Assays-by-Design primers and probes on an ABI 7900HT PCR machine, as previously described [16]. Dantu genotypes were infered from the rs186873296 *FREM3* allele, which is in strong linkage disequilibrium with the Dantu structural rearrangement [17]. For comparative purposes, we also genotyped participants for a range of other polymorphisms that have been reproducibly associated with protection from severe and complicated malaria in other studies. We typed for the common African form of G6PD deficiency, caused by the *G6PD* c.202T mutation [18], the blood group O mutation in *ABO* [19] and the rs4951074 allele in *ATP2B4* [5] using TaqMan SNP genotyping assays, and for the -α^3.7I^ deletional form of α^+^-thalassaemia by gap PCR [20].

### Statistical analysis

We considered three distinct outcomes, each capturing a different aspect of the parasitological and clinical progress of malaria infections: (a) whether infections progressed to reach the pre-defined treatment threshold of 500 parasites/µl; (b) whether or not participants received malaria treatment, for either clinical or parasitological reasons; (c) the time from inoculation to treatment in participants who did receive treatment. We made (a) the primary endpoint for this analysis because the hypothesis we were testing concerned *in vivo* parasite growth rather than susceptibility to symptoms. We conducted between-genotype comparisons both by univariate analysis and by multivariate analysis with adjustment for other malaria-protective genes, anti-schizont antibody concentration, and location of residence, of which the latter two were significantly associated with the same outcomes in an earlier analysis of the same cohort [21]. We used the Fisher’s exact test for differences in proportions and logistic regression for multivariate analyses. We used the Kruskal-Wallis and Dunn’s tests to investigate between-group differences in maximum parasitaemia. Finally, we compared time to treatment using Kaplan-Meier survival curves and Cox regression models for multivariate analyses. All statistical analyses were performed using R V3.6.2 [22].

### Ethics

The study was approved by both the KEMRI Scientific and Ethics Review Unit (protocol KEMRI/SERU/CGMR-C/029/3190) in Kenya, and the University of Oxford Tropical Research Ethics Committee (OxTREC; protocol 2-16) in the UK. The study was conducted based on good clinical practice (GCP) following the principles of the Declaration of Helsinki.

## Results

Of 161 volunteers recruited to the study, 19 were excluded, either because non-CHMI parasite strains were detected in post-CHMI samples (suggesting the presence of coincidental natural infections) (n=7), or because anti-malarial drugs were detected in pre-CHMI samples (n=12) [10]. After these exclusions, data from 142 individuals contributed to the current analysis. Genotyping revealed that 30 of these individuals were either heterozygous or homozygous for the Dantu allele. We did not get Dantu genotype data on one individual due to poor quality of the DNA sample, therefore Dantu genotype data on 141 out of the 142 participants was used in the downstream analysis.

### *The Dantu variant protects against* P. falciparum *growth* in vivo

While infections progressed to the point of reaching the pre-determined treatment threshold of 500 parasites/µl in 25/142 (17.6%) of all volunteers, the proportions varied markedly by Dantu genotype. None of the thirty (0.0%) Dantu-carriers reached this threshold in comparison to 25/111 (22.5%) of the non-Dantu individuals (p=0.01) (Table 1, Figure 1). The difference in the proportion of Dantu heterozygous (0/27; 0%) and non-Dantu individuals reaching the treatment threshold was strongly significant (p=0.004) but did not reach statistical significance in the case of Dantu homozygotes 0/3 (0.0%) because of the small number of individuals in this group.

**Table 1.** The proportion of participants reaching the pre-defining treatment threshold, by genotype category.

| Variant | Genotype | n/N | % | P-value overall | P-value Homozygous Reference vs Heterozygous | P-value Homozygous Reference vs Homozygous Derived |
| --- | --- | --- | --- | --- | --- | --- |
| Dantu<br>rs186873296 | non-Dantu (AA) | 25/111 | 22.5 | 0.01 | 0.004 | 1 |
|  | Heterozygous (AG) | 0/27 | 0.0 |  |  |  |
|  | Dantu Homozygous (GG) | 0/3 | 0.0 |  |  |  |
| G6PD +202<br>rs1050828 | Homozygous Reference (C/CC) | 20/110 | 18.2 | 0.505 | 0.739 | 0.463 |
|  | Heterozygous (CT) | 4/17 | 23.5 |  |  |  |
|  | Homozygous Derived (T/TT) | 1/15 | 6.7 |  |  |  |
| ABO<br>rs8176719 | non-O | 13/64 | 20.3 | 0.517 | 0.517 | - |
|  | O | 12/75 | 16.0 |  |  |  |
| α-thalassaemia | Homozygous Reference ($\alpha\alpha/\alpha\alpha$) | 6/48 | 12.5 | 0.408 | 0.328 | 0.29 |
| | Heterozygous ($-\alpha/\alpha\alpha$) | 14/70 | 20.0 | | | |
| | Homozygous Derived ($-\alpha/-\alpha$) | 5/21 | 23.8 | | | |
| ATP2B4<br>rs4951074 | Homozygous Reference (GG) | 11/62 | 17.7 | 0.876 | 1 | 0.749 |
|  | Heterozygous (AG) | 11/57 | 19.3 |  |  |  |
|  | Homozygous Derived (AA) | 3/23 | 13.0 |  |  |  |

n = the number of participants remaining untreated; N = the total number within the genotype category; $\alpha\alpha/\alpha\alpha$ no α-thalassaemia; $-\alpha/\alpha\alpha$ heterozygous α-thalassaemia; $-\alpha/-\alpha$ homozygous α-thalassaemia. \* For G6PD, male and female were combined as C/CC = normal (wild type hemizygous males and homozygous females), CT = carrier females and T/TT = G6PD deficient hemizygous males and homozygous females.

**Figure 1.**
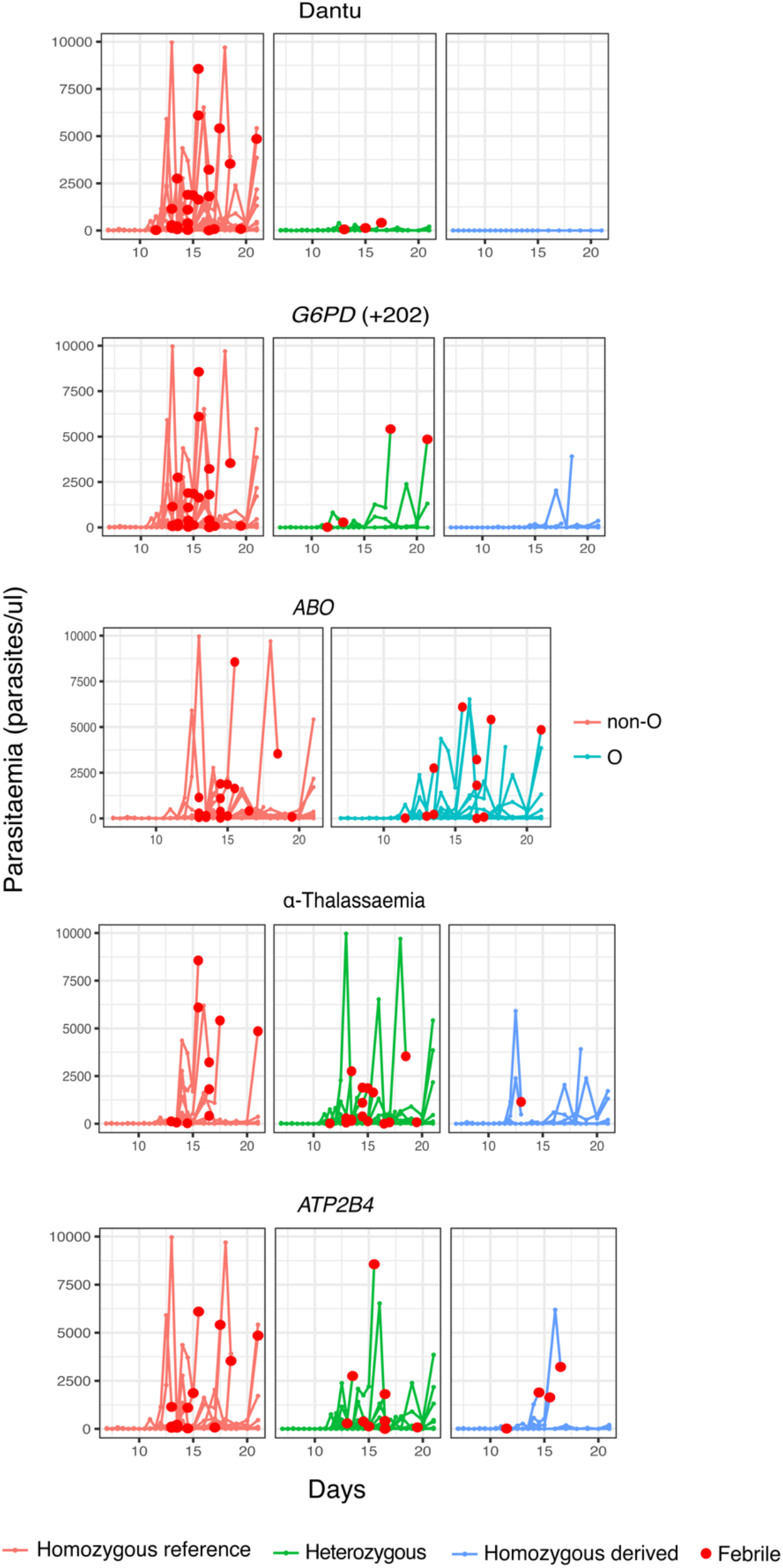
The impact of human genotype on parasite growth. Parasitaemia measured by quantitative PCR (y-axis) is shown over the duration of the study (x-axis). The red dots indicate individuals that were febrile and met treatment criteria.

### Fewer Dantu-carriers required malaria treatment

Although a threshold of 500 parasites/μl was considered the primary endpoint of the study and triggered the administration of anti-malarial treatment per-protocol, some participants developed symptoms, and were therefore treated, before reaching this threshold. Only one quarter (7/27; 25.9%) of the Dantu heterozygotes and none (0/3; 0.0%) of the Dantu homozygotes (Table 2, Supplementary Figure 1) received antimalarial treatment in comparison to 49/111 (44.1%) of the non-Dantu individuals. While this did not reach statistical significance on univariate analysis, on multivariate analysis, adjusting for other variants and confounders, Dantu-carrying subjects overall were found to have been administered anti-malarial treatment 79% less frequently (OR 0.21; 95% CI 0.05-0.71; p=0.021) than non-Dantu individuals (Table 3).

**Table 2.**
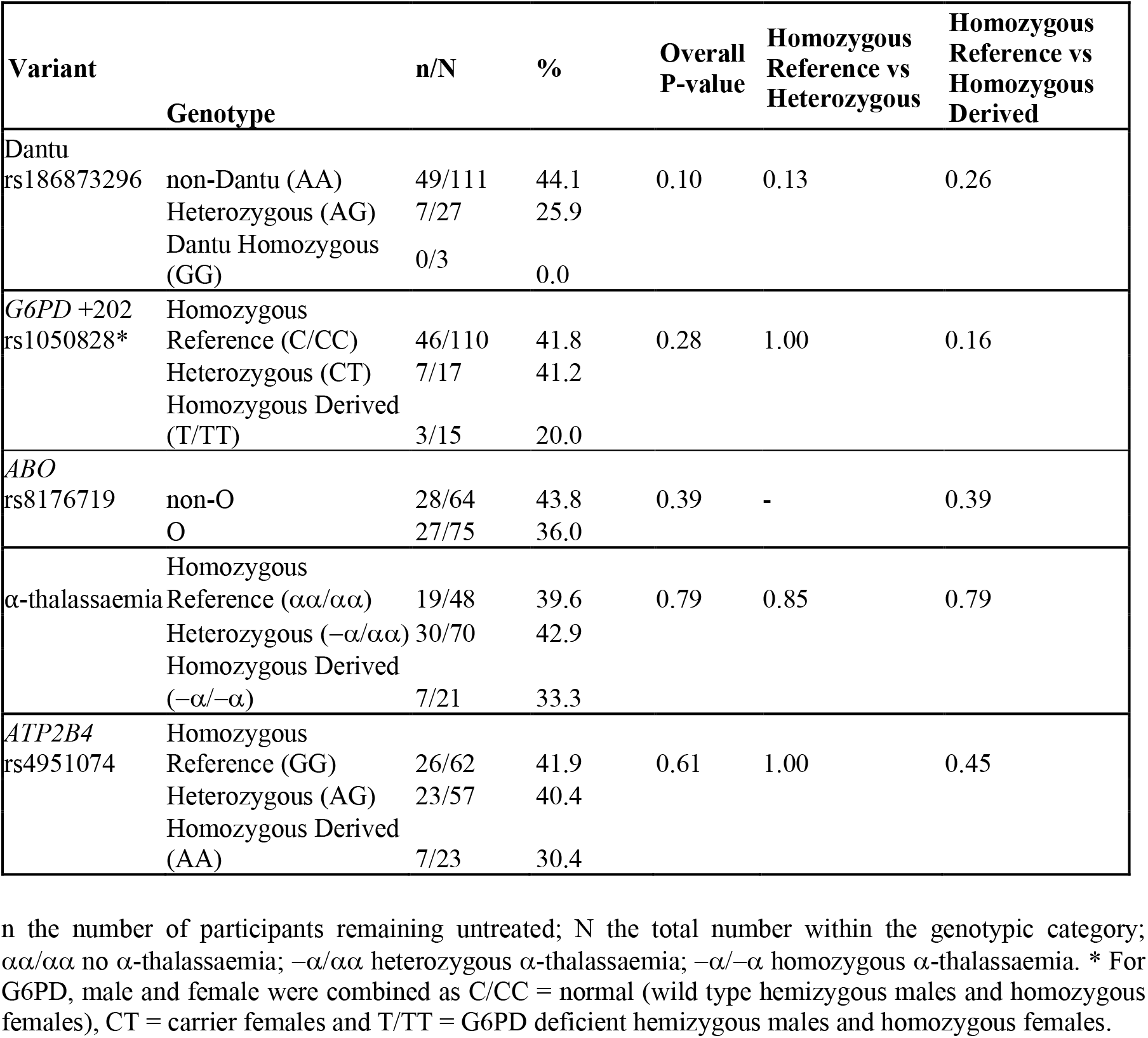
The numbers and frequencies of individuals who received treatment before day 21, by genotypic category.

**Table 3.**
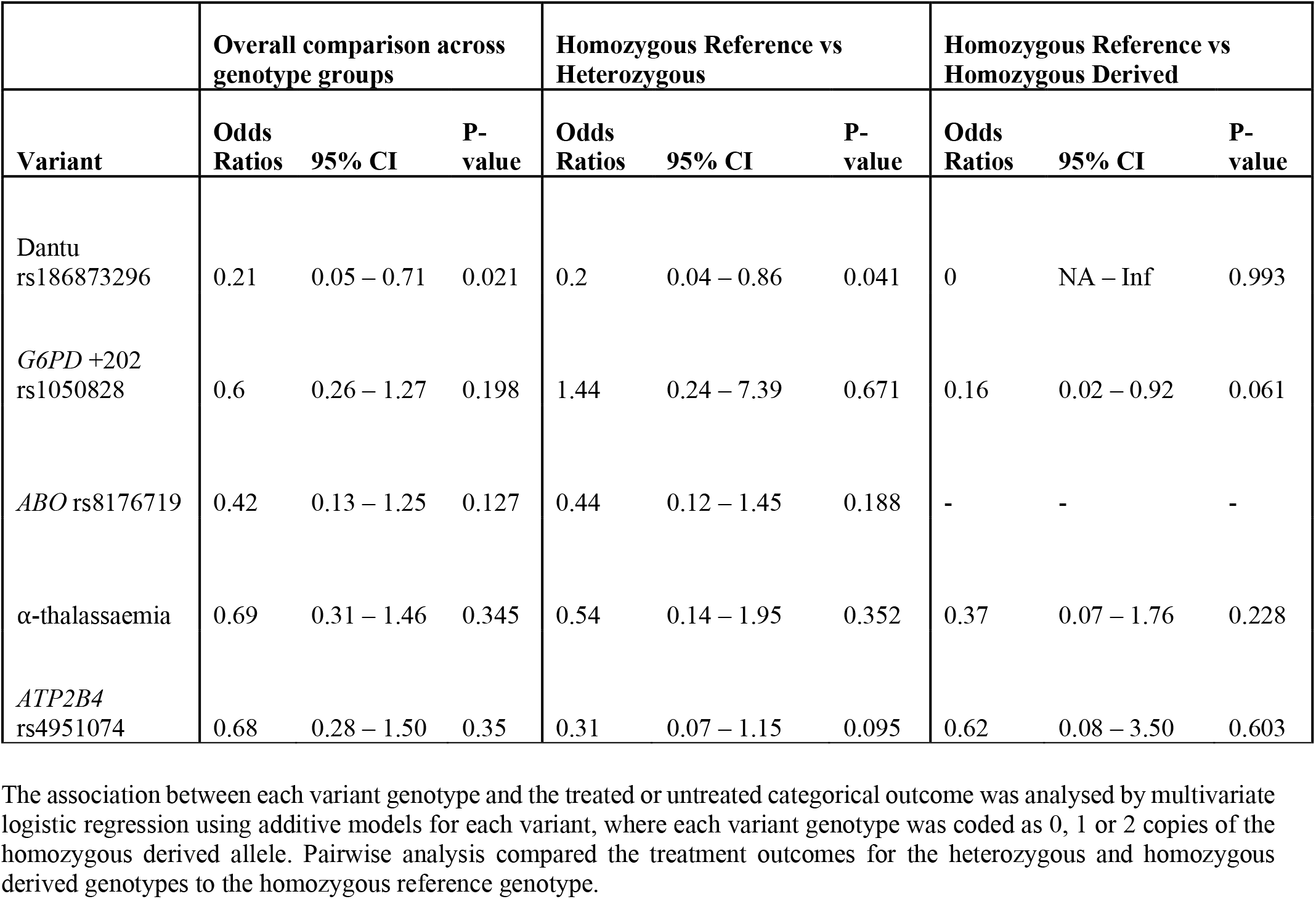
The impact of specific genes on treatment outcome.

### Peak parasitaemias were lower in Dantu-carrying participants

The proportion of participants who became PCR-positive at any parasitaemia was similar at 86/111 (77.5%) in the non-Dantu and 20/27 (74.1%) and 2/3 (66.7%) among Dantu heterozygotes and homozygotes respectively (P=0.745) (Supplementary Table 1). However, maximum parasitaemias were considerably higher in non-Dantu than in Dantu-carrying individuals. Peak parasitaemias reached 9,694 parasites/μl in the non-Dantu group in comparison with 411 parasites/μl in the Dantu heterozygotes and only 3 parasites/μl among the Dantu homozygotes (non-Dantu vs Dantu heterozygotes P=0.028; non-Dantu vs Dantu homozygotes P=0.141; and non-Dantu vs Dantu heterozygotes and homozygotes combined P= 0.009) (Supplementary Table 1, Supplementary Figure 2). Similarly, the median parasitaemia among those who did become PCR-positive was 112 parasites/μl in the non-Dantu, 13 parasites/μl in the Dantu heterozygous and 2 parasites/μl in the Dantu homozygous groups respectively, although these differences did not reach statistical significance (non-Dantu vs Dantu heterozygotes P=0.108; non-Dantu vs Dantu homozygotes P= 0.256; and non-Dantu vs combined Dantu heterozygotes and homozygotes P= 0.068) (Supplementary Table 1).

### Time to treatment was significantly longer in Dantu-carrying than non-Dantu individuals

Among the participants who did receive treatment, the time to treatment was significantly longer among Dantu-carrying than non-Dantu individuals. While the Hazard Ratio was 0.39 (CI 0.17-0.87; P=0.022) overall, a dose-dependent effect of the Dantu genotype was also seen, as none of the three Dantu homozygotes required treatment and the time to treatment was significantly longer in Dantu heterozygous than in non-Dantu individuals (HR=0.41, p=0.042) (Figure 2, Supplementary Figure 3).

**Figure 2.**
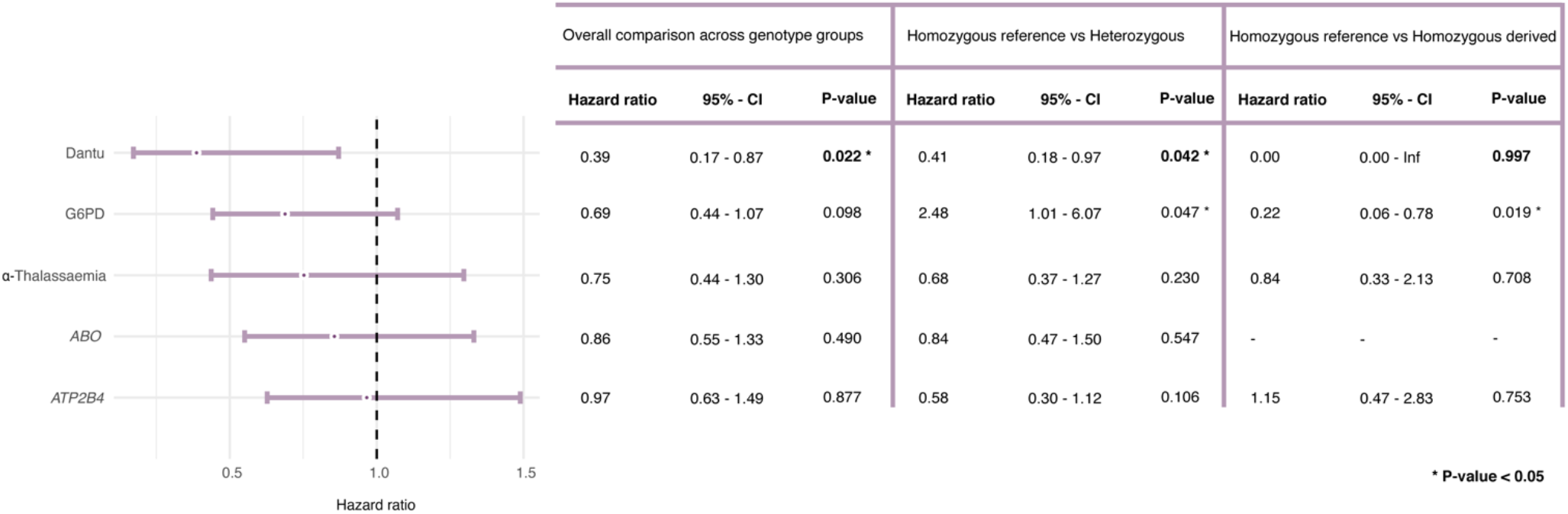
The impact of malaria-protective variants on time to treatment. The impact of each gene variant on the time to treatment was tested using multivariate Cox regression models, with each variant genotype coded as 0, 1, or 2 copies of the homozygous derived allele in an additive model, adjusting for the other four malaria-protective variants, anti-schizont antibody concentration and location of residence. Pairwise analysis compared the time to treatment in the heterozygous and homozygous derived genotypes to the homozygous reference genotype.

### No significant impacts were seen with regard to any of the outcomes under study for any of the remaining RBC polymorphisms

No significant differences were seen in any of the study outcomes between the different genotype groups for α-thalassaemia, blood group O, G6PD-deficiency or *ATP2B4* (Tables 1, 2 and 3, Figure 2).

## Discussion

Through the analysis of data from a CHMI study with PfSPZ Challenge injection conducted in Kenyan adults, we have shown that the Dantu blood group is associated with a high degree of protection against uncomplicated malaria infections. While more than 20% of Dantu-negative volunteers developed bloodstream malaria infections that reached a pre-defined threshold of 500 parasites/μl following controlled PfSPZ administration, this threshold was not reached by any of the 30 Dantu positive volunteers. This is the first time that Dantu has been shown to protect against early-stage malaria infections. Our study suggests that Dantu protects against severe and complicated malaria [7, 23] by preventing the disease from becoming established in its earliest phase. This is consistent with recent *in vitro* observations that have demonstrated a link between Dantu genotype and susceptibility to red blood cell invasion by *P. falciparum* merozoites, which we previously predicted would lead to reduced parasite growth *in vivo* [8]. That mechanistic study revealed that the Dantu genotype protects red blood cells from invasion by increasing their surface tension, which reduces the ability of merozoites to deform their surface and hence productively invade. Critically, this tension difference was greater in Dantu homozygotes than heterozygotes, as was the reduction in invasion efficiency, supporting a dose-dependent protective effect. This dose-dependency was similarly reflected in our current *in vivo* study, as while 44.1% of the Dantu negative volunteers developed symptoms that precipitated malaria treatment, this occurred in only 25.9% of Dantu heterozygous and in none of the three (0%) Dantu homozygous volunteers. Similarly, the maximum parasitaemia observed was considerably higher in the non-Dantu than in the Dantu heterozygous and homozygous participants.

In contrast to CHMI studies in malaria-naïve populations, where most participants typically develop clinical malaria [24], less than one-fifth of the participants in our current study reached a threshold of >500 parasites/μl. This is probably because most of the participants in our study were raised in malaria-endemic communities and will therefore have been partially immune to the disease. Indeed, in previous analyses of data from the same study we have shown that resistance was attributable to the degree of prior exposure, and was reflected by the titre of anti-schizont antibodies [21], while other correlates of protection included the antibody-dependent phagocytosis of both ring-infected and uninfected erythrocytes from parasite cultures [25] and the breadth of antibodies to *P. falciparum* Variant Surface Antigens [26]. As such, Dantu genotype was not the only factor at play in determining the clinical outcome in this study. Our multi-variate analysis was adjusted for these factors as previously [21], as well as other malaria-protective genetic variants, underscoring the protective effect conferred by Dantu.

The impact of Dantu on uncomplicated malaria was in stark contrast to that of the other genetic factors under study. Although consistent evidence has been found for protective effects against severe malaria by G6PD deficiency, blood group O, the rs4951074 allele in *ATP2B4* and α^+^-thalassaemia in numerous previous studies [5, 14], none of these polymorphisms had any significant impacts on any of the outcomes under investigation in our current study. This is probably because, unlike Dantu, none of these conditions have a clearly established impact on merozoite red cell invasion, but instead influence the progress of malaria once the disease has become established. For example, recent studies have shown that α^+^-thalassaemia has no effect on either red blood cell invasion [27] or the development of uncomplicated malaria [14, 23], but protects instead against the development of severe and complicated disease through mechanisms that include the reduced expression of red cell surface antigens that result in cytoadhesion [28, 29]. Similarly, it had no apparent impact on infectivity in a previous, smaller, CHMI study using PfSPZ Challenge, conducted in Tanzania [30].

In conclusion, our current study reveals the power of CHMI studies to deconvolute the malaria-protective effects of naturally occurring human genetic variants, establishes for the first time that the Dantu blood group provides strong protection against uncomplicated malaria, and emphasises the potential of Dantu-phenocopying interventions to limit *P. falciparum* growth *in vivo*.

## Data Availability

All data produced in the present work are contained in the manuscript

## Acknowledgements

The Kenya CHMI study was supported by an award from Wellcome (grant number 107499). SK was supported by a Training Fellowship (216444/Z/19/Z), TNW by a Senior Research Fellowship (202800/Z/16/Z) and JCR by an Investigator Award (220266/Z/20/Z), all from Wellcome. For the purpose of open access, the author has applied a CC BY public copyright licence to any Author Accepted Manuscript version arising from this submission.

## Author Information

**Centre for Geographic Medicine Research (Coast), Kenya Medical Research Institute-Wellcome Trust Research Programme, Kilifi, Kenya**

Silvia N Kariuki, Alex Macharia, Johnstone Makale, Wilfred Nyamu, Melissa C Kapulu, Philip Bejon & Thomas N Williams

**Centre for Tropical Medicine and Global Health, Nuffield Department of Medicine, University Oxford, Oxford, UK**

Melissa C Kapulu & Philip Bejon

**Cambridge Institute for Medical Research, University of Cambridge, Cambridge, UK**

Julian C. Rayner

**Department of Surgery and Cancer, Imperial College, London, UK**

Thomas N Williams

<u>Members of the CHMI-SIKA Study Team</u>

**Centre for Geographic Medicine Research (Coast), Kenya Medical Research Institute-Wellcome Trust Research Programme, Kilifi, Kenya**

Abdirahman I Abdi, Philip Bejon, Zaydah de Laurent, Mainga Hamaluba, Domtila Kimani, Rinter Kimathi, Kevin Marsh, Sam Kinyanjui, Khadija Said Mohammed, Moses Mosobo, Janet Musembi, Jennifer Musyoki, Michelle Muthui, Jedidah Mwacharo, Kennedy Mwai, Joyce M Ngoi, Omar Ngoto, Patricia Njuguna, Irene Nkumama, Francis Ndungu, Dennis Odera, Donwilliams Omuoyo, Faith Osier, Edward Otieno, Jimmy Shangala, James Tuju, Juliana Wambua, & Thomas N Williams

**Sanaria Inc**., **Rockville, MD, USA**

Yonas Abebe, Peter F Billingsley, Stephen L Hoffman, Eric R James, Thomas L Richie, & B Kim Lee Sim

**Centre for Tropical Medicine and Global Health, Nuffield Department of Medicine, University Oxford, Oxford, UK**

Sam Kinyanjui, & Kevin Marsh

**Department of Pathology, University of Cambridge, Cambridge, UK**

Peter C Bull

**Pwani University, P. O. Box 195-80108, Kilifi, Kenya**

Sam Kinyanjui & Cheryl Kivisi

**Epidemiology and Biostatistics Division, School of Public Health, University of the Witwatersrand, Johannesburg, South Africa**

Kennedy Mwai

**Centre for Infectious Diseases, Heidelberg University Hospital, Heidelberg, Germany**

Irene Nkumama, Dennis Odera & Faith Osier

**Centre for Clinical Research, Kenya Medical Research Institute, Kisumu, Kenya**

Bernhards Ogutu, Fredrick Olewe & John Ong’echa

**Center for Research in Therapeutic Sciences, Strathmore University, Nairobi, Kenya**

Bernhards Ogutu, Fredrick Olewe & John Ong’echa

