## Supplementary Information for "Controlled Human Malaria Infection reveals that the Dantu blood group variant provides high level protection against uncomplicated malaria"

**Supplementary Table 1.** Proportion of PCR-positive individuals across Dantu genotype groups among the CHMI participants

| Dantu rs186873296 genotype | Non-Dantu (n=86) | Dantu Heterozygous (n=20) | Dantu Homozygous (n=2) | P-value overall | P-value non-Dantu vs Dantu Heterozygous | P-value non-Dantu vs Dantu Homozygous |
|---|---|---|---|---|---|---|
| n/N (%) | 86/111 (77.5%) | 20/27 (74.1%) | 2/3 (66.7%) | 0.745 | 0.125 | 0.258 |
| Maximum Parasitaemia | 9694 | 411 | 3 | 0.028 | 0.020 | 0.141 |
| Median Parasitaemia | 112 | 13 | 2 | 0.144 | 0.108 | 0.256 |

The number and frequencies of individuals in each genotype category that were PCR-positive over the course of the CHMI study. n = the number of participants that were PCR-positive; N = the total number within the genotype category. Statistical comparisons of proportions of PCR-positive individuals across genotype groups and pairwise comparisons between genotype groups were performed by Fisher's Exact test. Statistical comparisons of maximum and median parasitaemia between genotype groups were performed using the Kruskal-Wallis test, and post-hoc Dunn's test for pairwise differences between the genotype groups.

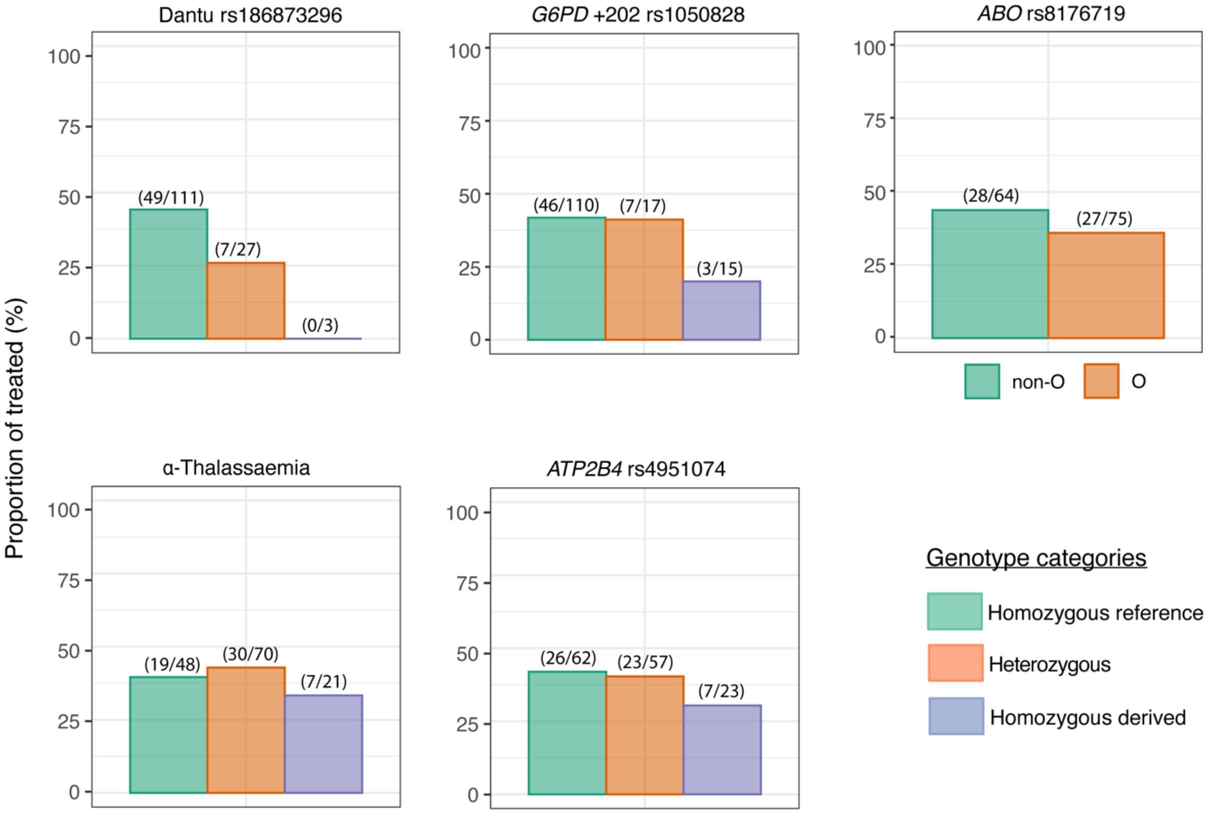


**Supplementary Figure 1**. The impact of each gene variant on treatment outcome. The proportion of individuals in each genotype category that were treated over the course of the CHMI study is shown on the y-axis, with the number of treated individuals out of the total number in each genotype given in parenthesis above the bar graphs.

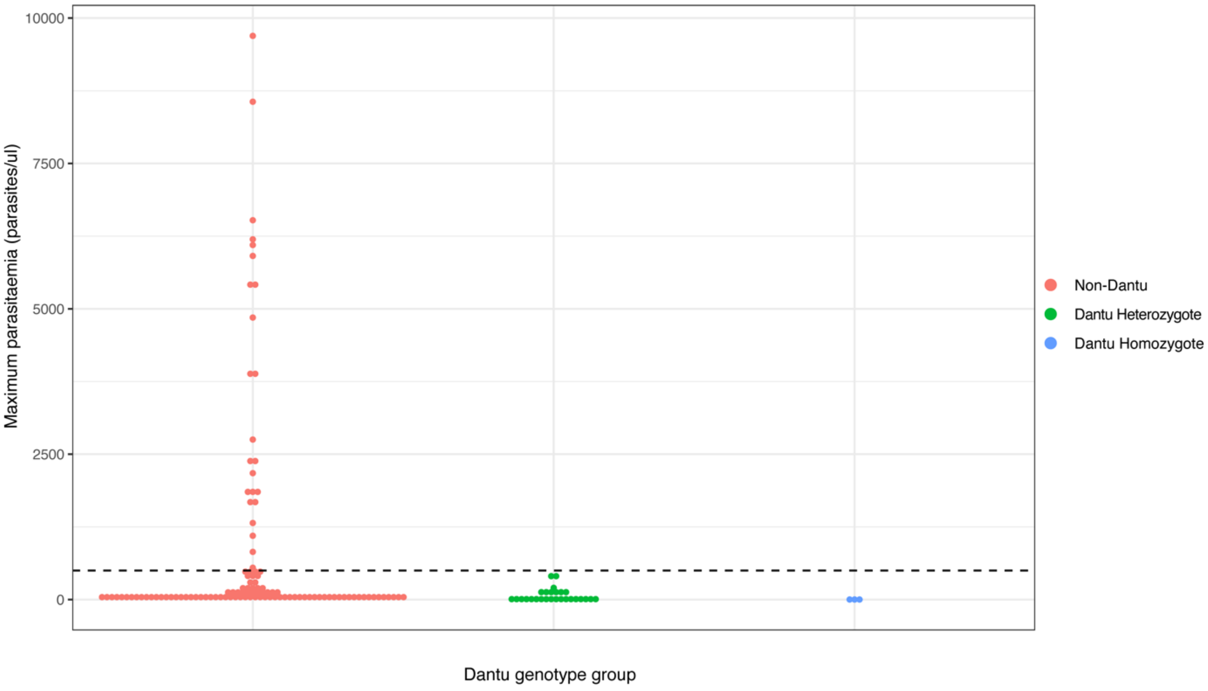


**Supplementary Figure 2.** Maximum parasitaemia values for individuals across Dantu genotype groups, with dashed line indicating the treatment threshold of 500 parasites/ul.

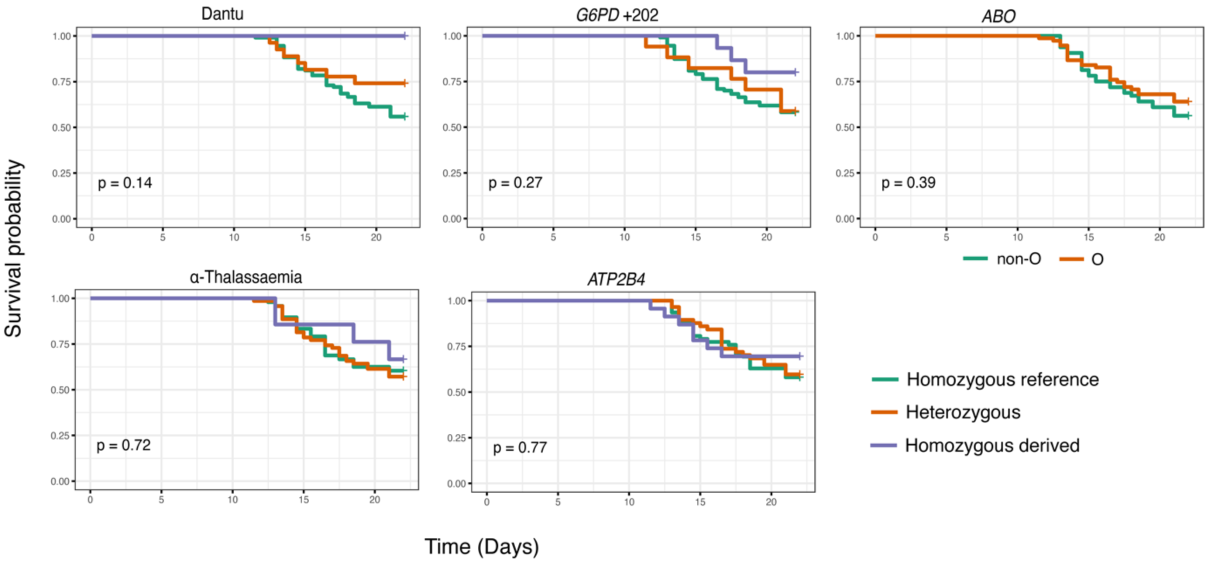


**Supplementary Figure 3**. Time to treatment survival analysis across malaria-protective genotypes. The time to treatment was analysed using Kaplan-Meier survival curves, with univariate comparisons across genotype groups performed using the Log-Rank test.
